# Transcatheter or Surgical Aortic Valve Replacement in Patients with Severe Aortic Stenosis and Small Aortic Annulus: A Randomized Clinical Trial

**DOI:** 10.1101/2023.09.28.23296187

**Authors:** Josep Rodés-Cabau, Henrique Ribeiro, Siamak Mohammadi, Vicenç Serra, Talal Al-Atassi, Andres Iniguez, Victoria Vilalta, Luis Nombela-Franco, Jose Ignacio Saez de Ibarra, Vincent Auffret, Jessica Forcillo, Lenard Conradi, Marina Urena, Cesar Moris, Antonio Muñoz-Garcia, Jean-Michel Paradis, Eric Dumont, Dimitri Kalavrouziotis, Pablo Maria Pomerantzeff, Vitor Emer Egypto Rosa, Mariana Pezzute Lopes, Carles Sureda, Victor Alfonso Jimenez Diaz, Carlos Giuliani, Marisa Avvedimento, Emilie Pelletier-Beaumont, Philippe Pibarot, the VIVA trial investigators

## Abstract

**BACKGROUND:** The optimal treatment in patients with severe aortic stenosis (AS) and small aortic annulus (SAA) remains to be determined. The objectives of this study were to compare the hemodynamic and clinical outcomes between transcatheter aortic valve replacement (TAVR) and surgical aortic valve replacement (SAVR) in patients with a SAA.

**METHODS:** Prospective multicenter international randomized trial performed in 15 university hospitals. Participants were 151 patients with severe AS and SAA (mean diameter <23 mm) were randomized (1:1) to TAVR (n=77) vs SAVR (n=74), The primary outcome was impaired valve hemodynamics (i.e. severe prosthesis patient mismatch [PPM] or moderate-severe aortic regurgitation [AR]) at 60 days as evaluated by Doppler-echocardiography and analyzed in a central echocardiography core laboratory. Clinical events were secondary outcomes.

**RESULTS:** The mean age of the participants was 75±5 years, with 93 of women, a median STS of 2.5 (1.7-3.3)%, and a mean annulus diameter of 21.1±1.2 mm.

**CONCLUSIONS:** This trial will provide clinicians with scientific evidence to determine if population with smaller aortic anatomy in the setting of severe AS maybe better suited to TAVR compared with SAVR.

**TRIAL REGISTRATION:** Clinicaltrials.gov: NCT03383445

## INTRODUCTION

Degenerative aortic stenosis (AS) remains the most common valvular disorder affecting western countries. A significant proportion of AS patients harbor a small aortic annulus (SAA), particularly among the women population.^1^ The treatment of this group of patients remains challenging, with a higher incidence of suboptimal hemodynamic results and its potential deleterious clinical impact following aortic valve replacemen.^1-3^ In the last decade, transcatheter aortic valve replacement (TAVR) has become an effective alternative to surgical aortic valve replacement (SAVR) for the treatment of AS. Current guidelines recommend both treatment options for treating elderly patients with AS, but with a preference for SAVR as a default option in younger and lower risk patients.^4,5^ However, aortic annular size, prosthetic valve hemodynamic results, and gender factors are not taken into consideration in current guideline recommendations.

Data from observational studies and sub-studies from randomized trials suggest superior prosthetic valve hemodynamics following TAVR (vs. SAVR) in patients with SAA.^6-8^ Additionally, several studies have reported improved outcomes associated with TAVR in women and patients of Asian descent,^9-12^ both of which constitute populations with a high prevalence of SAA. However, women have often been largely under-represented in heart valve trials,^13^ and no randomized studies exist regarding the optimal treatment of women with severe AS and SAA. The objective of this multicenter randomized trial was to compare the hemodynamic and clinical outcomes of TAVR vs. SAVR for the treatment of patients with AS and SAA.

## METHODS

### Trial design and patients

The VIVA trial (Clinicaltrials.gov: NCT03383445) was a prospective randomized controlled trial (investigator initiated). The study was conducted in 15 centers in Canada, Europe, and Brazil, and included elderly (≥65 years old) patients with severe AS and SAA considered eligible for either SAVR or TAVR by the multidisciplinary heart team of each participating center. Exclusion criteria included the presence of aortic root dilatation >45 mm, coronary artery disease not treatable by percutaneous coronary intervention or coronary artery bypass grafting, SYNTAX score >32, significant concomitant mitral or tricuspid valve disease, or prior aortic valve surgery. Clinical and exclusion criteria are listed in Table 1. Patients were initially screened on the basis of an aortic annulus diameter <21 mm as evaluated by transthoracic echocardiography, and the final inclusion of the patient was based on the presence of a mean aortic annular diameter <23 mm and a minimal diameter ≤21.5 mm as evaluated by contrast computed tomography (CT). Severe aortic stenosis was defined as (i) a jet velocity ≥ 4.0 m/s or mean gradient ≥ 40 mmHg or velocity ratio <0.25, and aortic valve area ≤ 1.0 cm^2^ or aortic valve area index ≤ 0.6 cm^2^/m^2^; or (ii) mean gradient >30 mmHg and aortic valve area ≤ 1.0 cm^2^ or aortic valve area index ≤ 0.6 cm^2^/m^2^, and >1200 Agatston units for women or >2000 Agatston units for men as determined by non-contrast CT. The study was approved by the Ethics Committee of each participating center, and all patients provided signed informed consent for participating in the trial (trial protocol, including the changes in inclusion criteria and reasons, are reported as Supplemental material).

**Table 1.** Study inclusion and exclusion criteria.

|  |
| --- |
| **Inclusion criteria** |
| Patients $\geq 65$ years-old diagnosed with severe AS (defined as: jet velocity $\geq 4.0$ m/s or mean gradient $\geq 40$ mmHg or velocity ratio $< 0.25$ AND aortic valve area $\leq 1.0$ cm<sup>2</sup> or aortic valve area index $\leq 0.6$ cm<sup>2</sup>/m<sup>2</sup>;<br>OR<br>mean gradient $> 30$ mmHg AND aortic valve area $\leq 1.0$ cm<sup>2</sup> or aortic valve area index $\leq 0.6$ cm<sup>2</sup>/m<sup>2</sup> AND $> 1200$ Agatston units for women or $> 2000$ Agatston units for men as determined by non-contrast CT).<br>-Small aortic annulus defined as a mean aortic annulus diameter $< 23$ mm and a minimal aortic annulus diameter of $\leq 21.5$ mm as measured by 3D-computed tomography (CT) and/or 3D-transesophageal echocardiography (TEE). |
| **Exclusion criteria** |
| -Prohibitive surgical risk as determined by the Heart Team<br>-Porcelain aorta<br>-Aortic root dilatation $> 45$ mm<br>-Coronary artery disease (CAD) not treatable by percutaneous coronary intervention (PCI) or coronary artery bypass grafting (CABG), or SYNTAX score $> 32$ (in the absence of prior revascularization) or severe left main disease<br>-Non-calcific aortic stenosis<br>-Severe mitral regurgitation<br>-Moderate-to-severe tricuspid regurgitation requiring surgical repair<br>-Prior surgical valve in aortic position |

Patients were randomly assigned in a 1:1 ratio to receive TAVR or SAVR. Random block sizes of 10 were used to conceal treatment allocation and implementation was assessed by an independent statistician (not related to the study). Randomization was centralized, and performed using the electronic case-report form (eCRF) system. Participants and site staff were unblinded to the treatment assigned.

### Study procedures

The TAVR procedures were performed according to routine clinical practice in each participating center, and only newer generation valve systems were used in the study, including the balloon-expandable SAPIEN 3/ULTRA valve (Edwards Lifesciences, Irvine, CA), and the self-expandable Evolut R/PRO/PRO+ valve (Medtronic, Minneapolis, MN), and the Acurate Neo/Neo2 valve (Boston Scientific, Boston, MA). Prosthesis valve sizing was based on CT measurements and followed the recommendations from the manufacturers. The SAVR procedures were performed according to the standards of the surgical team of each participating center, and all surgical prosthetic valves approved for clinical use were allowed in the study. Also, the decision regarding aortic root enlargement was left to the criteria of the surgeon responsible for the intervention.

### Outcomes

The primary efficacy outcome was impaired valve hemodynamics defined as the occurrence of severe prosthesis-patient mismatch (PPM) and/or moderate-severe aortic regurgitation (AR) as evaluated by Doppler-echocardiography at 60 days. Secondary end points included the mean transvalvular gradient at 60 days, and clinical endpoints (death, stroke, major or life-threatening bleeding, new-onset atrial fibrillation, permanent pacemaker implantation, cardiac rehospitalization) at 30 days and at follow-up. Patients were followed at 30, 60 days, 1 year, and yearly thereafter up to 5 years. Clinical events were defined according to VARC-2 criteria (14). The functional status (NYHA class) and quality of life (as determined by the Kansas City Cardiomyopathy Questionnaire [KCCQ]) were evaluated at baseline and at each follow-up visit. There were no changes to trial outcomes after the trial commenced.

Echocardiographic findings (for echocardiography examinations at 60 days post-intervention) were evaluated in a central Echocardiography Core Laboratory (Core Lab of the Quebec Heart & Lung Institute, Quebec City, Canada), blinded to the allocated treatment. Severe PPM was defined per-protocol as an indexed effective orifice area (EOA) ≤0.65 cm^2^/m^2^ (14). Severe PPM was also assessed according to VARC-3 criteria, defined as an indexed EOA ≤0.65 cm^2^/m^2^ for patients with BMI <30 kg/m^2^ and as an indexed EOA ≤0.55 cm^2^/m^2^ for patient with BMI ≥30 kg/m^2^ (15).

### Statistical analysis

Based on the results of a sub-study from the PARTNER 1 trial (7), the expected rate of severe PPM in the TAVR group was evaluated at 19%, with a rate of moderate-severe AR of 3% (overall incidence of the primary endpoint of 22%). In the surgical group, the expected rate of severe PPM was estimated at 38%, without any anticipated moderate-severe AR (7). A total of 130 patients per group were calculated to provide 80% power to detect significant differences in the composite rate of PPM or moderate-severe AR between groups with a p value <0.05, and the final sample size of the study was estimated at 300 patients (150 patients per group). The study started in August 2017, with a progressive inclusion of participating centers within the 1^st^ two years of the study. Once all centers were activated and enrolling patients, the COVID-19 pandemic appeared and this translated into a drastic reduction in the enrollment rate in all centers. Additionally, the results of 2 randomized trials showing the non-inferiority or superiority of TAVR for treating aortic stenosis in elderly patients were published during the execution of the study (16, 17), further impending the enrolment of patients after the COVID-19 pandemic. It was finally decided to prematurely stop study enrolment in April 2023, when 156 patients (52% of the estimated sample size) had been included in the study.

The primary and secondary end point analyses were performed on an intention-to-treat basis. A secondary “as treated” analysis including only those patients who finally received the allocated treatment was also performed. Qualitative variables were expressed as percentages and quantitative variables as mean (standard deviation) or median (interquartile range) depending on variable distribution. Comparison of numerical variables was performed using the Student’s t test or Wilcoxon rank sum test. Survival curves for time-to-event variables were performed using Kaplan-Meier estimates, and the log rank test was used for group comparisons. Event rates of clinical events at 30 days and at follow-up were summarized as mean difference (95% CI). A p value <0.05 was considered significant. The analyses were performed with SAS (version 9.4).

## RESULTS

The flowchart of the study population is shown in **Figure 1**. A total of 156 patients were randomized, and 79 and 77 patients were allocated to TAVR and SAVR, respectively. Of these, 5 patients were excluded (consent withdrawn before treatment=2; intended use of a transcatheter valve non-approved for the study=1; significant mitral regurgitation requiring surgical intervention=1; not eligibility for the study after second CT evaluation=1), and a total of 77 and 74 patients were finally included in the TAVR and SAVR groups, respectively. All patients but 1 received the allocated treatment in the TAVR group (1 patient in the TAVR group finally underwent SAVR), and 2 patients in the SAVR group finally had a TAVR procedure (these 3 patients remained in the study and were included in the allocated groups for the intention-to-treat analyses).

**Figure 1.**
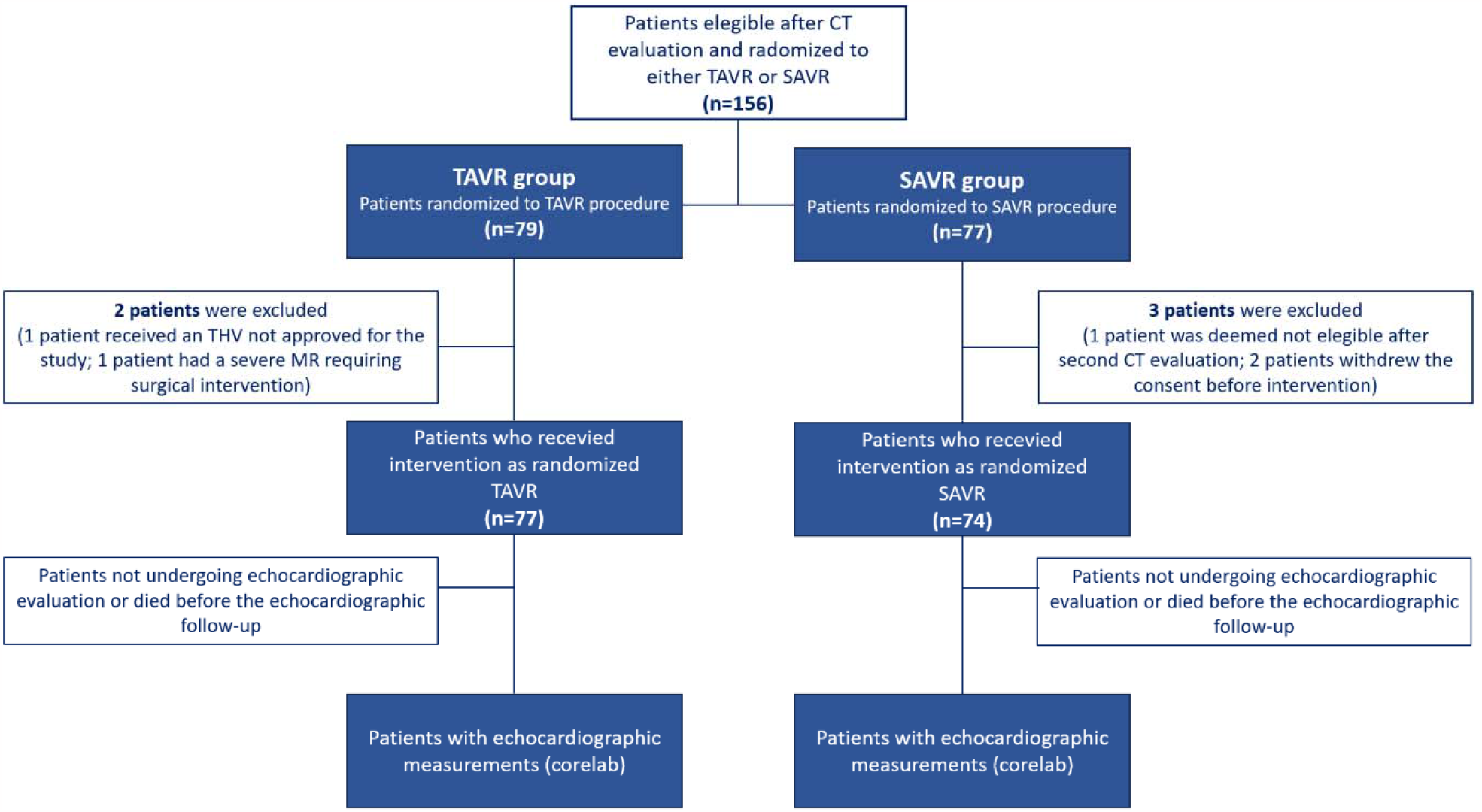
Study flowchart.

The main baseline and procedural characteristics of the study population are shown in **Table 2**. The mean age and median STS of the study population were 75.5 (5.0) years and 2.5 (1.67-3.28) %, respectively, and the vast majority of patients (93%) were women. The mean and minimal aortic annular diameters as evaluated by CT were of 21.1 (1.2) mm and 18.6 (1.5) mm, respectively.

**Table 2.** Baseline Characteristics of the Study Population.

|  | **n=151** |
| --- | --- |
| **Clinical characteristics** |  |
| Age, years | 75.5±5.1 |
| Women | 140 (92.7%) |
| BMI, kg/m<sup>2</sup> | 28.2±5.3 |
| BSA, m<sup>2</sup> | 1.71±0.19 |
| NYHA class III-IV | 47 (31.1%) |
| STS, % | 2.5 (1.7-3.3) |
| Diabetes | 45 (29.8) |
| Hypertension | 123 (81.5%) |
| Atrial fibrillation | 20 (13.2%) |
| Renal insufficiency* | 51 (33.8%) |
| COPD | 21 (13.9%) |
| Coronary artery disease** | 31 (20.5%) |
| Prior pacemaker | 4 (2.6%) |
| Mean aortic annulus diameter (computed tomography), mm | 21.1±1.2 |
| Minimal aortic annulus diameter (computed tomography), mm | 18.6±1.5 |
| Aortic annulus area (computed tomography), mm<sup>2</sup> | 345±40 |
| Aortic annulus perimeter (computed tomography), mm | 66.8±5.7 |
| **Echocardiographic data** |  |
| Left ventricular ejection fraction, % | 62±8 |
| Mean aortic gradient, mm Hg | 48±17 |
| Maximal aortic gradient, mm Hg | 80±24 |
| Aortic valve area, cm<sup>2</sup> | 0.71±0.29 |
| Moderate-severe aortic insufficiency¶ | 9/147 (6.1%) |

\*eGFR<60 ml/min/m<sup>2</sup> \*\*Presence of significant coronary lesions (≥70% diameter stenosis) requiring treatment or prior CABG/PCI ¶The suboptimal quality of echocardiographic images precluded any evaluation of aortic regurgitation in 4 patients Values are expressed as mean±SD, n (%) or median interquartile range; STS= Society of Thoracic Surgeons predicted risk of mortality;

## DISCUSSION

The VIVA trial is the first randomized trial in the field of AS field dedicated to SAA patients, including a vast majority (93%) of women. The study has been designed to evaluate the hemodynamic performance (incidence of severe PPM and ≥ moderate AR) between TAVR and SAVR in patients with severe AS and small aortic annuli. A secondary objective was to compare clinical outcomes (death, stroke, major or life threatening bleeding) in TAVR and SAVR recipients harboring small aortic annuli.

### Importance of knowledge to be gained

Results of this trial will make an important contribution to the management of patients with small aortic annulus by determining whether TAVR will be hemodynamically superior to SAVR for treating patients with severe AS and a small aortic annulus. The strict anatomical criteria applied in a prospective manner and using CT parameters in this trial will translated into the inclusion of a population with true SAA, including a mean aortic annulus diameter and area of ∼21 mm and <350 mm^2^, respectively. This anatomical feature presented a clear sex-related association, with the vast majority of patients being women, and this was in accordance with prior studies including AS patients with SAA.^1,18,19^ This could be largely explained by the small BSA in women compared to men,^20^ but specific sex-specific differences beyond BSA parameters cannot be excluded. Some previous studies reported worse clinical outcomes in women (vs. men) undergoing SAVR, including higher mortality rates and a higher incidence of periprocedural complications.^21,22^ Indeed, some studies including a recent meta-analysis showed improved outcomes with TAVR (vs. SAVR) at early and midterm follow-up among women.^23-25^

### Limitations

Although the study is a prospective, randomized trial, the sample size of the study was limited and the possibility of a type II error for some clinical variables cannot be excluded. Also, there will be no clinical event adjudication committee for this trial. The early termination of the trial could have an impact on the primary endpoint results of the study.

## Conclusions

There is limited clinical trial data on hemodynamic performance between TAVR and SAVR in patient with severe AS and small aortic annuli. This trial will provide clinicians with scientific evidence to determine if population with smaller aortic anatomy in the setting of severe AS maybe better suited to TAVR compared with SAVR.

## Data Availability

The data referred to in the manuscript will be available to other authors upon reasonable request and only for the purpose of patient-level meta-analyses.

## ACKNOWLEDGMENTS

Dr. Rodés-Cabau holds the Research Chair “Fondation Famille Jacques Larivière” for the Development of Structural Heart Disease Interventions (Laval University, Quebec City, Canada).

## FUNDING

Investigator initiated trial, with financial support from the Research Chair “Fondation Famille Jacques Larivière” for the Development of Structural Heart Disease Interventions (Laval University, Quebec City, Canada); grant from the Foundation of the Quebec Heart & Lung Institute (Quebec City, Canada); and grant from the Research Center of the Quebec Heart & Lung Institute (Quebec City, Canada).

## CONFLICT OF INTEREST

Dr. Rodés-Cabau has received institutional research grants from and is consultant for Edwards Lifesciences and Medtronic. Dr. Nombela-Franco has received consulting fees from Edwards Lifesciences. Dr Conradi is a member of the Advisory Board of Medtronic, Abbott, JenaValve, and Neovase; and has received consulting fees from Edwards Lifesciences, Boston Scientific, New Valve Technology, and MicroInterventions. Dr Moris received fees as a proctor from Medtronic and Boston Scientific. Pibarot has received institutional research grants from Edwards Lifesciences, Medtronic, Pi-Cardia, and Novartis. The rest of authors do not report any significant conflict of interest with respect to the content of this article.

## Notes

### Clinical Trial

NCT03383445

### Author Declarations

Comite ethique de la recherche Institut universitaire de cardiologie et de pneumologie de Quebec-Universite Laval Ethical approval - Ethical approval was given Comissao Nacional de Etica Em Pesquisa - Plataforma Brasil - Ethical approval was given Comite de etica de investigacion con medicamentos y comision de proyectos de investigacion del hospital universitary Vall Hebron - Ethical approval was given Ottawa Health Science Network Research Ethics Board- Ethical approval was given Comite de Etica de Investigacion con medicamentos de Galicia - Ethical approval was given Comite de Etica de la investigacion con medicamentos del Principado de Asturias- Ethical approval was given Comite de Etica en Investigacion Clinica Hospital Clinico San Carlos- Ethical approval was given Comite etica de la investigacion de las Islas Baleares- Ethical approval was given Comite de Protection des Personnes Ile de France II - Ethical approval was given Ethik-Kommission Der Arztekammer Hambur- Ethical approval was given Comite de Etica de la investigaciOn con medicamentos del Principado de Asturias- Ethical approval was given ComitE de Etica en InvestigaciOn ClInica Hospital Universitario Virgen de la Victoria- Ethical approval was given

### Summary of Updates

This version of the manuscript has been revised to update the results section.

